# Effects of online tDCS and hf-tRNS on reading performance in children and adolescents with developmental dyslexia: a study protocol for a within-subject, randomized, double-blind, and sham-controlled trial

**DOI:** 10.1101/2023.07.25.23292956

**Authors:** Andrea Battisti, Giulia Lazzaro, Cristiana Varuzza, Stefano Vicari, Deny Menghini

## Abstract

**Background:** Developmental Dyslexia (DD) is a brain-based developmental disorder characterized by severe impairments in the acquisition of fluent and/or accurate reading. The extensive data on the neurobiology of DD have increased interest in *brain-directed* approaches. Transcranial direct current stimulation (tDCS) has been proposed as a non-invasive method to address reading difficulties in individuals with DD. While overall positive outcomes have been observed, the results remain heterogeneous. To enhance the current findings, various modalities have been employed, including manipulation of electrode montages, experimental designs, and targeting different brain regions. However, no studies have yet investigated the comparative effects of two different types of transcranial electrical stimulation, namely tDCS and transcranial random noise stimulation (tRNS), on reading abilities in children and adolescents with DD.

**Methods:** The present study will apply a within-subject, randomized, double-blind, and sham-controlled design. The aim of the present study is to investigate the effects of tDCS and tRNS on reading abilities in children and adolescents with DD. Participants will undergo three conditions, each separated by a one-week interval: (A) a single session of active tDCS; (B) a single session of active tRNS; and (C) a single session of sham (tDCS or sham tRNS). The order of the sessions will be counter-matched among participants. Left anodal/right cathodal tDCS and bilateral tRNS will be applied over the temporo-parietal regions for a duration of 20 minutes. The assessment of text, word, and non-word reading measures will be conducted immediately before and during each stimulation sessions. Safety, tolerability and blinding parameters will be assessed using a questionnaire.

**Results:** Our hypothesis is that tRNS will demonstrate comparable effectiveness to tDCS in improving text, word, and non-word reading measures compared to the sham conditions. Additionally, we anticipate that tRNS will exhibit a similar safety profile to tDCS.

**Conclusion:** This study has the potential to contribute novel insights into the effectiveness of tRNS, a newly-wave tES method that has not yet been explored in children and adolescents with DD. Furthermore, findings may lay the groundwork for further investigations involving multiple tRNS sessions.

**Trial registration:** The study has been registered with ClinicalTrials.gov under the identifier NCT05832060.

## INTRODUCTION

Reading acquisition is a critical milestone in human development – particularly in modern literate societies, as it serves as a foundation for appropriate educational, professional, and social functioning.

Developmental Dyslexia (DD) poses a significant challenge in achieving fluent and accurate reading skills, despite adequate cognitive abilities and educational opportunities (1). DD is widely acknowledged as one of the most common neurodevelopmental disorders in childhood, significantly affecting the overall well-being and mental health of individuals (2,3). Furthermore, the adverse effects of DD persist into adulthood, leading to long-term consequences (4).

Extensive research has focused on investigating the neurocognitive architecture of reading, revealing the involvement of a widespread brain network responsible for different reading processes (5). This intricately interconnected reading network includes the left dorsal temporo-parietal cortex – encompassing the posterior superior temporal gyrus, the supramarginal gyrus, and the angular gyrus, which plays a crucial role in grapheme-phoneme conversion. Additionally, the left ventral occipito-temporal cortex progressively specializes in orthographic coding during literacy development and facilitates rapid word recognition (6).

There is consistent evidence demonstrating reduced activation in the left temporo-parietal and the left occipito-temporal reading network among individuals with DD (7), which opens up possibilities for the application of *brain-directed* interventions. In fact, when considering behavioural interventions for DD, effective and long-lasting effects are lacking (8,9).

Transcranial electrical stimulation (tES) techniques have been proposed as a non-invasively means to target atypical brain functioning in individuals with DD, with the aim of improving specific aspects of reading, such as text accuracy or speed, word recognition, and non-word decoding (5).

tES involves the application of a weak, low-intensity (0.5-2 mA) electrical current through electrodes placed on the scalp over specific cortical areas. These electrodes are typically covered by square or rectangular sponges soaked in saline solution (10). The primary mechanism of action involves modulating neuronal membrane excitability below the threshold for generating action potentials (11,12).

Among tES techniques, tDCS is the most widely used method in the paediatric population. It is a polarity-dependent technique, consisting of a continuous electrical current delivered via two types of electrodes: anode (positive current) or cathode (negative current) (10). Generally, anodal tDCS induces depolarisation of the membrane potential via increasing the excitability of the brain areas, whereas cathodal tDCS induces opposite effects therefore inhibiting cortical excitability (11–13). Neurophysiological studies suggest that tDCS can induce neuroplasticity aftereffects, leading to LTP-like processes through Ca2+ and NMDA receptor-dependent plasticity (11,14,15).

Several studies have demonstrated the effects of tDCS, either alone or in combination with reading training, on reading abilities in children and adolescents with DD (16–22). While overall positive outcomes have been observed, the results still show heterogeneity in terms of specific reading aspects improved (e.g., non-word speed v*s* text reading accuracy). Over time, different approaches have been employed to enhance the current results, including the manipulation of electrode montages (e.g., bilateral: left anodal/right cathodal, right anodal/left cathodal; unilateral: left anodal, left cathodal), utilization of different experimental designs (one-session vs. multi-sessions; between-subjects vs. within-subjects; stand-alone vs. combined with cognitive training), and targeting various brain regions (inferior frontal gyrus, inferior parietal lobe, posterior middle temporal gyrus, supramarginal gyrus, superior temporal gyrus, temporo-parietal cortex, temporo-parietal junction, V5/MT).

Apart from tDCS, transcranial random noise stimulation (tRNS) is another tES technique that is gaining attention, although its application in children and adolescents is still limited (23–26).

tRNS is a polarity-independent technique that involves the application of alternating electrical current at random intensities (e.g., ±0.5 mA) and frequencies (i.e., full spectrum: 0.1-640 Hz; low-frequency range: 0.1-100 Hz; high-frequency range: 100-640 Hz) (27). The exact neurophysiological mechanisms underlying the effects of tRNS are still unclear and subject to debate (28). However, two main hypotheses have been advanced. First, a phenomenon called *stochastic resonance* has been theorised as responsible for tRNS-induced effects. When an optimal level of noise is added to a weak, subthreshold, noise signal (i.e., brain oscillatory activity), the sum of the signals will exceed the threshold at some point (29). The amplification of subthreshold oscillatory brain activity, which in turn reduces the amount of endogenous noise, improves the signal-to-noise ratio, leading to enhanced perception or cognitive performance.

Secondly, *in vitro* and pre-clinical studies suggested that tRNS can induce neuroplasticity processes via LTP through the shortening of hyperpolarization phase and the repetitive re-opening of Na^+^ channels (30–32).

In comparison to tDCS, the effectiveness of tRNS in improving reading abilities has been less extensively explored. A study by Rufener et al. (24) compared the effects of a single session of both tRNS and a different tES technique, transcranial alternating current stimulation (tACS), targeting the bilateral auditory cortex. This study aimed to investigate the online effects of tES on auditory phoneme processing in adolescents (Study 1) and adults (Study 2) with DD. While a positive effect emerged in adolescents only during tACS compared with tRNS and sham condition, a significant improvement was found in adults during tRNS compared with the sham condition. Furthermore, a very recent study by Bertoni et al. (33) found that a single session of active tRNS applied over the bilateral posterior parietal cortex, significantly increased word reading in typical adult readers compared to the sham condition. These findings suggest that, in addition to tDCS, tRNS is a promising and effective modality for improving reading and related processes, such as auditory phoneme processing, in children and adolescents with DD.

Nevertheless, to the best of our knowledge, the direct effects of tRNS on reading in individuals with DD have not been tested. Additionally, although tDCS and tRNS have been compared in populations with and without neurodevelopmental disorders [e.g., ADHD: (25,26); healthy adults: (34,35)], their effects have not been directly compared in individuals with DD. Regarding children and adolescents with ADHD, tRNS has shown to be more effective than tDCS in improving ADHD symptoms, working memory, and processing speed (25,26). Concerning healthy adults, tRNS has demonstrated better results compared to tDCS in tasks involving working memory and divergent/convergent thinking (34,35).

Both tDCS and tRNS are considered highly promising tES techniques for the paediatric population due to their favourable safety profile, high tolerability, versatility, ease of use, and low cost (25,36– 41). Interestingly, concerning safety and feasibility, both neurophysiological and behavioural studies [respectively, (25,35,36)] showed that tRNS has higher skin perception thresholds, lower response rates to adverse events, and more effective blinding, making it a preferable option for the paediatric population.

The current study aims to investigate whether tRNS is effective in improving reading abilities and whether it may outperform tDCS in specific reading aspects. The main goal is to compare the effects of a single online session of tRNS, tDCS, and sham stimulation on text, word, and non-word reading accuracy and speed in children and adolescents with DD. This proof-of-concept study will also assess the safety, tolerability, and blinding parameters of both tES techniques and compare them.

If tRNS proves to be at least as effective as tDCS in improving reading outcomes, these results would provide valuable insights for the development of multi-session tRNS protocols as a potential treatment approach for individuals with DD.

## METHODS AND ANALYSIS

### Ethical Committee

The local research ethics committee (process number 2639_OPBG_2021) has granted ethical approval for this study, which has been registered on ClinicalTrials.gov (ID: NCT05832060). The study will be conducted in compliance with the Declaration of Helsinki. The protocol adheres to the SPIRIT guidelines (Standard Protocol Items: Recommendations for Interventional Trials) and is prepared using the SPIRIT 2013 Checklist (42).

### Participants

Participants will be recruited during the daily clinical activities of the I.Re.Ne Lab (Innovation and Rehabilitation in Neurodevelopment Lab) by neuropsychiatrists, psychologists, and speech therapists from the Child and Adolescent Neuropsychiatry Unit of the Bambino Gesù Children’s Hospital in Rome. When appropriate, participants will be selected from a large database managed by the Head of the Unit (S.V.), which includes several hundred patients evaluated in accordance with the good clinical practices of international guidelines for neurodevelopmental disorders. Research assistants will contact selected participants via phone and email to provide information about the ongoing project and assess their interest in participating. All participants and their parents will receive comprehensive instructions regarding the experimental procedures and objectives. The principal investigator will obtain written consent before participants are enrolled in the study.

### Clinical Eligibility

The inclusion criteria will be as follows: (1) participants of both genders, who have been diagnosed with DD based on the Diagnostic and Statistical Manual of Mental Disorders, Fifth Edition [DSM-5; (1)] and national recommendations. Their assessment will be conducted by experienced psychologists and neuropsychiatrists using “gold standard” assessment tools (43–46) and considering their developmental history; (2) intelligence quotient (IQ) ≥ 85; (3) age between 8 years and 13 years and 11 months; and (4) normal or corrected-to-normal vision.

The exclusion criteria will be as follows: (1) presence of another primary psychiatric diagnosis (e.g., depression, anxiety), autism, or ADHD; (2) personal history of neurological/medical/genetic diseases; (3) personal or first-degree relatives’ history of epilepsy; (4) receiving any concomitant treatment for DD during the enrolment in the project; and (5) currently receiving any Central Nervous System (CNS)-active drug treatment.

### Assessment

The assessment will be conducted in a dimly lit room.

Participants’ cognitive level will be evaluated using non-verbal cognitive tests such as the Coloured Progressive Matrices [CPM; (47)] and the Standard Progressive Matrices [SPM; (48)]. Alternatively, a multi-componential cognitive test such as the Wechsler Intelligence Scale for Children, 4^th^ edition [WISC-IV; (49)] may be used.

To meet the criteria for DD, participants’ accuracy or speed levels must be at least 1.5 standard deviations (SD) below the normative data for their school-age group, and these difficulties should significantly interfere with their school and daily functioning.

All participants will be assessed for emotional-behavioural symptoms using the Child Behaviour Checklist [CBCL; (50)] and the Conners Parent Rating Scale [CPRS; (51)]. The Kiddie Schedule for Affective Disorders and Schizophrenia for DSM-5 (52) will be utilized to exclude neuropsychiatric comorbidities based on the inclusion/exclusion criteria.

Working memory will be assessed using verbal and visual-spatial n-back tasks. Participants will be required to indicate if a pronounced letter (verbal n-back) corresponds to the last spoken letter or whether a coloured box (visual-spatial n-back) moves to the same previous position. If accuracy will achieve 80%, the difficulty will increase and participants will be asked to remember not the last letter spoken or the last position shown, but the second-to-last (2-back), and so on (3-back, 4-back, etc.). An n-back performance index will be calculated and considered for each task (22,53).

In the phoneme blending task, participants will be instructed to combine individual phoneme sounds to form non-words. The number of accurately blended phonemes (Phonemes_Acc_) and the total time taken in seconds for each non-word (Phonemes_Time_) will be recorded and considered for analysis.

In the Rapid Automatized Naming task (RAN) for letters (RAN_Letters_) or colours (RAN_Colours_), participants will be asked to name aloud the letters or colours presented in lists as quickly and accurately as possible. Total time in seconds will be considered for each task.

To assess attentional shift abilities, an adapted version (54) of the Posner Cueing Task will be used. Participants will be seated in front of a monitor screen at eye level and instructed to focus on a central dot. Two circles will be simultaneously presented on the left and right sides of the fixation point. A vertical arrow above the circles will serve as a spatial cue, and a dot in the centre of one of the circles will act as the target. Participants will be instructed to maintain fixation on the central dot throughout the trial. Each trial will begin with the onset of the fixation point. After 504 ms, the two circles will appear, followed by the cue after another 504 ms. Subsequently, the target will appear inside one of the circles. Two different stimulus onset asynchronies (SOAs) will be used: 136 ms and 238 ms, presented randomly. On valid trials, the target will appear in the circle indicated by the cue, while on invalid trials, the target will appear in the opposite circle of the cue. Neutral trials will have both circles cued (one arrow on each circle), and the target will be randomly presented in one of the circles. Participants will be instructed to react as quickly as possible to the target onset by pressing the spacebar on a computer keyboard, and their reaction times and errors will be recorded for analysis. The maximum response time allowed will be 1 second.

### Study design

The study will employ a within-subject, randomized, double-blind, and sham-controlled design. Clinical eligibility will be assessed on Day 0 (Baseline). Participants will be exposed to three conditions with a one-week intersession interval (Day 1, Day 2, Day 3, as shown in Figure 1): (A) a single active tDCS session; (B) a single active tRNS session; and (C) a single sham tDCS or sham tRNS session. The ordering of conditions will be counter-matched among participants. After recruitment, participants will be allocated to one of six possible combinations of conditions (i.e., ABC, ACB, BAC, BCA, CBA or CAB). Stratified randomization will be performed by an independent researcher immediately after the participant completes the screening evaluation. This process will ensure assignment masking. Stratified randomization will use the minimal sufficient balancing method to prevent imbalances in baseline characteristics and will take into account participants’ demographics (e.g., age, IQ, gender) and the severity of DD (SD below the average in text, word and non-word reading tasks) (43–46). The randomization information will be retained by an independent researcher until the data collection has been completed.

**Figure 1.**
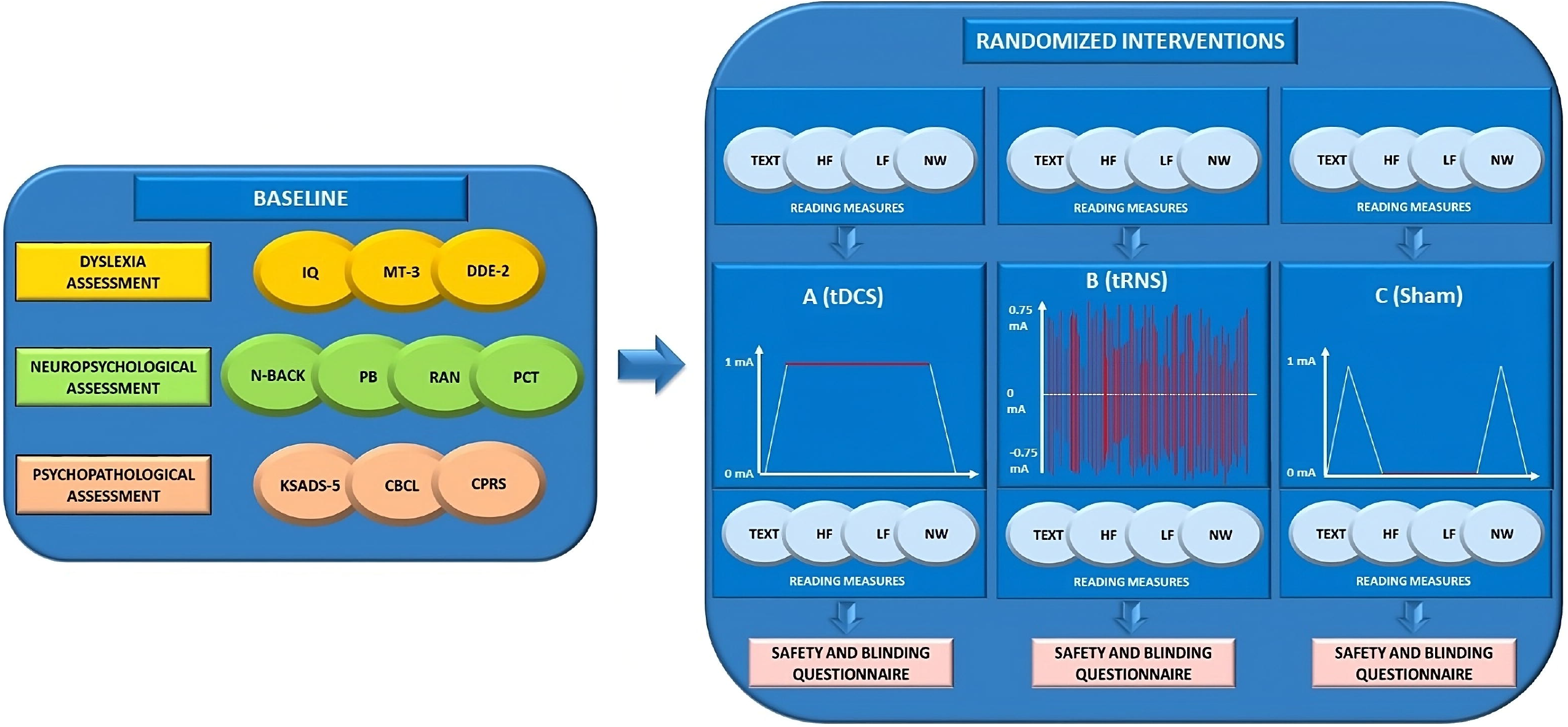
Overview of the study design. IQ, intelligence quotient; MT-3, (44,45); DDE-2, (43); N-BACK, (22,53); PB, Phoneme Blending; RAN, Rapid Automatized Naming Task; PCT, Posner Cueing Task (54); KSADS-5, Kiddie Schedule for Affective Disorders and Schizophrenia for DSM-5 (52); CBCL, Child Behaviour Checklist (50); CPRS, Conners Parent Rating Scale (51); TEXT, texts reading task; HF, high-frequency words reading task; LF, low-frequency words reading task; NW, non-words reading task; tDCS, transcranial direct current stimulation; tRNS, transcranial random noise stimulation; mA, milliamperes.

The principal investigator will dispose of an emergency code for each participant that can be revealed in case of a compelling need, such as a serious adverse event that requires awareness of current interventions to handle the participant’s condition.

Immediately before each session (Day 1, Day 2, and Day 3) and during the maximum peak of tES effects [approximately 10 min after the start of stimulation; (55)], participants will undergo an assessment that will include text, words and non-words reading. The order of the tasks administered will be randomized and counter-balanced across participants. Each session will last approximately 35 minutes. This includes 10 minutes for the reading tasks conducted before the stimulation session, 20 minutes for the stimulation session (active or sham) combined with concurrent reading tasks (initiated after 10 minutes of stimulation), and 5 minutes for the administration of the safety and blinding questionnaire after the stimulation session.

At the end of the final session (Day 3), participants will be informed of their assigned condition (i.e., ABC, ACB, BAC, BCA, CBA or CAB) and provided with a brief feedback on the results that emerged from each session.

### Interventions

To ensure protocol adherence and minimize variability, the experimenter responsible for delivering the interventions will complete a structured checklist before each stimulation session. This checklist will include participants’ information, details of the procedures to be applied, and dose parameters. The checklist is adapted from Antal et al. (56). See Supplementary Materials.

#### Active tES conditions

##### Transcranial direct current stimulation (tDCS)

According to the International 10–20 System, the anodal electrode will be placed over the left temporo-parietal region (TP7/P7), while the cathodal electrode will be placed over the contralateral temporo-parietal region (TP8/P8). Consistent with our previous tDCS protocols in developmental age (16–22), in the active condition a constant current at 1 mA intensity will be delivered for 20 min, with a density of 0.04 mA/cm^2^, preceded by 30 s of rump up (0 mA to 1 mA) and succeeded by 30 s of rump down (1mA to 0 mA). Direct current will be delivered by a battery driven, direct current stimulator (BrainStim stimulation by E.M.S. s.r.l.— Bologna, Italy) via a pair of identical, circular (25 cm^2^) saline-soaked (0.90 mol) sponge electrodes.

##### Transcranial random noise stimulation (tRNS)

During the active tRNS session, participants will receive 0.75 mA (±0.375 mA) of tRNS (100–500 Hz) to their temporo-parietal regions via 2 saline-soaked (0.90 mol) 25 cm^2^ circular sponges, placed over TP7/P7 and TP8/P8 based on the International 10–20 System. The current will be delivered by a BrainStim stimulator (E.M.S. s.r.l.; Bologna, Italy) for 20 min per session, as in previous tRNS protocols (23,25,57–61). The impedance of the electrodes will be checked before and during the application of tRNS to ensure that it remains below 10 kW.

#### Sham tES conditions

To control for potential placebo effects, participants during the sham condition will undergo the same procedures as those in the active conditions (active tDCS, active tRNS). This includes using the same electrode positioning and tRNS/tDCS equipment turn-on time (30 s). Apart from this short stimulation, during the sham condition participants will not receive the real stimulation (0 mA) for the rest of the session.

Among all participants, half of them will undergo a sham tDCS session (with the same electrode positioning and tDCS equipment turn-on time), while the other half will undergo a sham tRNS session (with the same electrode positioning and tRNS equipment turn-on time).

All participants, their families and the evaluators will be blinded to the stimulation conditions.

### Reading tasks

Four different reading tasks (text – TEXT; high-frequency words – HF; low-frequency words – LF; non-words – NW) will be presented to the participants, who will have to read aloud as quickly and accurately as possible.

To avoid the repetition effect, multiple versions of each reading task will be administered. This will include different versions presented at baseline (before stimulation) and during each stimulation session, resulting in a total of six versions across the study. The purpose of this approach is to minimize the impact of task familiarity. To control for the influence of fatigue, the order of the reading tasks will be counterbalanced between the three conditions (tDCS, tRNS, and sham).

Regarding reading task characteristics, each TEXT will be composed of more than 400 syllables, each list of HF will be composed of 10 trisyllabic and 10 bisyllabic high-frequency words, each list of LF will be composed of 10 trisyllabic and 10 bisyllabic low-frequency words, and each list of NW will be composed of 10 trisyllabic and 10 bisyllabic non-words.

For each reading task (TEXT, HF, LF, and NW), both reading speed and accuracy will be measured and considered. Reading speed will be quantified using the syllables per second ratio, calculated by dividing the total number of syllables pronounced by the total time taken to complete the reading task (in seconds). In terms of accuracy, errors such as omissions, substitutions, and letter additions will be considered as one-point errors. Auto-corrections will be considered as one-point errors only for TEXT, while for the remaining (HF, LF, and NW), they will not be treated as errors. The percentage of accuracy will be computed by dividing the number of correctly read stimuli by the total number of stimuli presented, multiplied by 100.

To ensure that the six versions of each reading task (TEXT, HF, LF, and NW) are comparable in terms of difficulty for both accuracy and speed, a pilot study was conducted with a group of typically developing readers. This preliminary investigation helped determine that the task versions will be appropriately matched in difficulty across the different conditions (see Supplementary Materials).

### Outcomes

The primary outcome of the study will be the changes in TEXT reading accuracy during the active tDCS and active tRNS sessions compared to the sham session (tDCS/tRNS), in comparison to the baseline measurement on Day 0.

Similarly, the secondary outcomes of the study will examine changes in TEXT reading speed, as well as changes in the accuracy and speed of the other reading tasks (HF, LF, and NW) during the active tDCS and active tRNS sessions compared to the sham session (tDCS/tRNS), in comparison to the baseline measurement on Day 0.

#### Safety, tolerability, and blinding assessment

Adverse effects will be assessed using a questionnaire adapted from Antal and colleagues (56). Participants will be asked to complete the questionnaire after each session. The questionnaire includes items related to potential adverse effects such as headache, tingling, skin redness, neck pain, scalp pain, itching, drowsiness, difficulty concentrating, burning sensation, and acute mood change. In addition, other information regarding when the adverse event occurred, its duration, and where the discomfort is located will be recorded. Participants will quantify the severity of adverse effects as follows: (0) absent; (1) mild; (2) moderate; and (3) severe.

The questionnaire will also inquire about participants’ perception of whether they received active stimulation or not. For more details, see Supplementary Materials (Paragraph: *Safety and tolerability questionnaire*).

### Sample size

The sample size is calculated on the primary outcome by a priori analysis in G*Power, version 3.1.9.7 (The G*Power Team, Düsseldorf, Germany). Based on the study design and assumptions, the estimated results suggest that participants who receive a single session of active tDCS or tRNS will demonstrate an improvement in TEXT reading accuracy compared to their baseline performance. On the other hand, participants who receive a single session of sham tDCS or sham tRNS are not expected to show a significant change in their performance compared to baseline. While the design of this project has never been employed in DD, we will refer to a study with a similar design (one-session, within-subject, sham-controlled experiment, comparing baseline vs. tDCS vs. tRNS vs. sham) assessing other cognitive functions (34).

Based on these previous results, to be cautious and conservative, we estimate a medium effect size (f) of 0.25. With an estimated f = 0.25, α value = 0.05 (i.e., probability of false positives of 5%), and β = 0.80 (i.e., at least 80% power), the sample size is 24 as calculated using a Repeated Measures-Analysis of Variance (RM-ANOVA) model with four within factors (baseline, active tDCS, active tRNS, and sham).

### Statistical Analyses and Expected Results

The Shapiro–Wilk test will be used to test the normality of the data and Levene’s test for the homogeneity of variances. When data will be normally distributed and the assumption of homogeneity will not be violated, parametric analyses will be computed. When one assumption will not be met, non-parametric tests will be conducted or a log-transformation of the distribution will be applied. Sphericity will be verified by Mauchly’s sphericity test and when not met, Greenhouse– Geisser correction will be applied.

Chi-Square analyses will be used to compare the groups on demographic and safety and blinding measures (categorical variables).

A preliminary analysis to test the effect of the four repetitions (Day 0, Day 1, Day 2, and Day 3) of the reading tasks (accuracy and speed of TEXT, HF, LF, and NW) will be conducted.

RM-ANOVA will be used to compare reading measures (TEXT, HF, LF, and NW accuracy and speed), separately, with conditions (Day 0, A, B, C) as a within-subjects factor.

Post hoc comparisons will be assessed using Tukey’s honest significance test.

Partial eta squares (ηp^2^) will be used as measures of effect sizes. We hypothesize that:

a. tRNS will be at least as effective as tDCS in improving TEXT reading accuracy compared with sham condition (primary outcome);
b. tRNS and tDCS will improve performance in the remaining reading tasks (TEXT reading speed and HF, LF, and NW reading accuracy and speed) compared with sham condition;
c. tRNS and tDCS will be as safe as sham condition;
d. the blinding will be kept across the conditions.

## DISCUSSION

Over the last decades, several interventions have been developed to improve reading skills in children and adolescents with DD (8,9), although with poor results in term of efficacy and long-term achievements. The growing understanding of the neurobiology of reading in individuals with and without DD has led to the exploration of brain-directed methods as a potential new frontier in the treatment of DD.

Promising findings on the application of tES to improve reading in individuals with DD are already available and represent the rationale for this project (62). However, there is still a need to bridge the gap and reduce the variability in results, thereby optimizing neuromodulation protocols. This includes identifying the most effective technique to employ.

The present study aims to compare two tES techniques already used in paediatrics, namely tDCS and tRNS, to investigate (i) which is more effective in improving reading abilities in children and adolescents with DD, and (ii) which is the most suitable in terms of safety and tolerability.

tDCS is definitely the most widely used technique to ameliorate reading performance in DD. One of the reasons is that numerous investigations have allowed to determine reliable tDCS parameters in terms of intensity and duration to achieve plastic after-effects, particularly by combining tDCS with transcranial magnetic stimulation at the level of the motor cortex (27). Moreover, tDCS polarity-dependent nature resulted suitable for left-hemisphere lateralization of reading [i.e., hypoactivation of a left hemisphere brain network and hyperactivation of contralateral homologous regions; for a review, see (5)], considering the possibility to simultaneously manipulate excitation/inhibition balance, via anodal/cathodal montage over target brain areas. Consistently, we will employ the left anodal/right cathodal montage accordingly to several studies that demonstrated the superiority of this montage in improving reading performance (62). Specifically, we will target the temporo-parietal regions that are crucially involved in reading, particularly in the process of letter-to-sound conversion (63–65).

Conversely, tRNS is a polarity-independent form of tES that employs the same electrode arrangement as tDCS to boost neuronal activity, but uses both electrodes to increase cortical excitability (27,30).

Previous findings have shown stronger effects of tRNS compared to tDCS on language and learning abilities, including mathematical skills (66), as well as on other cognitive (25,26,34,35) and perceptual processes (37).

Our selection of tES parameters, including intensity, density, duration, and frequency, will be based on previous research demonstrating the safety and beneficial effects of these parameters on reading performance (16–22) and cognitive tasks (23,57,60,67,68).

Concerning tDCS, we will employ a tDCS set-up that delivers a constant current at 1 mA intensity for 20 min. This configuration aligns with previous studies that have demonstrated the safety and tolerability of this approach in paediatric populations (16–22).

As for tRNS, we will apply a current intensity of 75% of 1 mA. Concerning the frequency band, tRNS can administer current in three distinct frequency ranges: a full-frequency range (typically spanning from 0.1 to 640 Hz), a low-frequency range (typically ranging from 0.1 to 100 Hz), and a high-frequency range (typically ranging from 101 to 640 Hz). Our decision to use the high-frequency tRNS set-up is founded on evidence demonstrating that only at 100–500 Hz of frequency tRNS provides consistent and long-lasting cortical excitability (30).

With respect to the experimental design, the choice of a within-subjects design is motivated by the aim to reduce inter-subject variability, which is well-known in the effects of tES [for a review, see (69)]. Within-subjects designs offer comparable accuracy to between-subjects designs and enhance statistical power even with a smaller number of participants (70–72) and suppressing inter-subject variability (73,74). In between-subject designs, there is a greater risk that stable factors specific to each participant may influence responses to tES conditions. In particular, it has been demonstrated that individuals anatomical differences (e.g., skull thickness, cortex morphology, and gyrification) could influence the electric field reaching the brain (75–77), and consequently modulate the behavioural effects of tES. Moreover, several studies showed that individual’s genotype seems to modulate synaptic plasticity and neurotransmitters expression, somehow affecting the effects of tES (78). Furthermore, it has been found that major demographic characteristics such as gender (79,80) and age (81–83) could contribute to the inter-subject variability in the effects of tES.

Among the main purposes of the present study is to compare the safety and tolerability profile of tDCS and tRNS in paediatrics, in order to verify which intervention is safer and more suitable for use as standard-of-care intervention for children and adolescents with DD. While tES has been increasingly applied in developmental age, there are limited studies that have systematically investigated the safety and tolerability of tES in children and adolescents [(38,81–83); for a review, see (39)], overall promoting tES as a safe method as for adults. Furthermore, while evidence suggests that tRNS is a safe and potentially less perceptible technique compared to tDCS (25,35,36), only one study has directly compared the safety and tolerability of the two techniques in children and adolescents (25), demonstrating that tRNS is less perceptible and potentially preferable in paediatrics.

## Limitations

While emphasizing the novelty of our approach and its rationale, we would also like to discuss some potential limitations.

First, to reduce the number of stimulation sessions per participant and streamline the study, we will not administer a sham session for both tDCS and tRNS. In fact, half of the participants will receive a sham tDCS session, while the other half will receive a sham tRNS session.

Second, the absence of neurophysiological measures, such as EEG, represent a limitation of the study. These measures could provide valuable insights into the underlying neurophysiological mechanisms of tDCS and tRNS and help interpret the observed behavioural outcomes. Given the partial understanding of the neurophysiological effects of tES techniques, incorporating neurophysiological measures alongside behavioural assessments would be beneficial for future studies.

## CONCLUSIONS

In conclusion, the present study could provide new insights regarding the effectiveness of a newly-wave tES method never used in children and adolescents with DD, the tRNS, potentially paving the way for further studies involving multiple tRNS sessions.

Of importance, this project will employ an experimental design that keeps the study burden for DD participants at a minimum, while providing key steps to understand which tES technique may be most effective for multi-session treatment applications.

The opportunity to compare tDCS and tRNS in the same experimental setting, investigating whether differ in modulating reading performance and in terms of safety and tolerability, will be a valuable step forward to identify brain-directed treatments for DD.

## Data Availability

All data produced in the present study are available upon reasonable request to the authors

## DECLARATIONS

### Compliance with ethics guidelines

The local research ethics committee (process number 2639_OPBG_2021) provided ethical approval for this study, which was registered at ClinicalTrials.gov (ID: NCT05832060). This study will be conducted in accordance with the Declaration of Helsinki.

### Consent for publication

Not applicable.

### Data availability statement

Not applicable.

### Authors’ contributions

DM, SV, and AB designed the study. AB, GL and DM drafted the manuscript, with support of CV. DM, SV and AB supervised the pilot study. All authors contributed to the article and approved the submitted version.

## Acknowledgments

We would like to thank in advance all the children and adolescents who will take part in the study and their parents. We also thank Claudia Varone for her contribution to the project.

