## Supplementary Materials for "Effects of online tDCS and hf-tRNS on reading performance in children and adolescents with developmental dyslexia: a study protocol for a within-subject, randomized, double-blind, and sham-controlled trial"

*Author details:*

*Andrea Battisti*, Child & Adolescent Neuropsychiatry Unit, Bambino Gesù Children’s Hospital, IRCCS, 00146 Rome, Italy; Department of Human Science, LUMSA University, 00193 Rome, Italy;

*Giulia Lazzaro*, Child & Adolescent Neuropsychiatry Unit, Bambino Gesù Children’s Hospital, IRCCS, 00146 Rome, Italy;

*Cristiana Varuzza*, Child & Adolescent Neuropsychiatry Unit, Bambino Gesù Children’s Hospital, IRCCS, 00146 Rome, Italy;

*Stefano Vicari*, Child & Adolescent Neuropsychiatry Unit, Bambino Gesù Children’s Hospital, IRCCS, 00146 Rome, Italy; Department of Life Science and Public Health, Catholic University of the Sacred Heart, 00168 Rome, Italy;

*Deny Menghini**, Child & Adolescent Neuropsychiatry Unit, Bambino Gesù Children’s Hospital, IRCCS, 00146 Rome, Italy.

* Correspondence:

Deny Menghini

ORCID ID: 0000-0002-3459-2601

### SUPPLEMENTARY MATERIALS

#### Checklist adapted from Antal et al (2017)

*A structured checklist increases the reproducibility of studies, minimizes deviations from a given protocol and diminishes variability. A structured checklist is thus the recommended procedure for enhancing reliability and comparability in publications of TES experiments/trials.*

1) Participant information

- Age:
- Gender:
- Handedness:
- Head size (distance in cm: inion – nasion, ear to ear distance):
- *Previous experience with TES (additional information of potential relevance):*
- Medication (Depending on the type of study an even more precise documentation may be necessary, measurement of drug levels may be considered), label and dose:

Within last hours

Within last days

Within last months

- Caffeine consumption (cups) (indicate the best currently relevant estimate):

Within last 12 h

Average within last months

- Nicotine consumption (cigarettes per day) (indicate the best currently relevant estimate):

Within last 4 h (*half life of Nicotine: 2 h*)

Within last 48 h (*half life metabolite cotinine: 10–37 h*)

- Alcohol consumption (drinks) (indicate the best currently relevant estimate):

Within last 24 h

Average with last months (how many months?)

- Drugs (e.g. marijuana) consumption (to be specified):

(for comparability important that unit is given and comparable measures are noted)

- Hormonal/menstrual cycle of female subjects
- *In case of patients non-neuropsychiatric comorbidities:*

2) Procedures applied, Dose parameters *(sufficient information about the stimulation parameters should be provided in order to replicate or model the stimulation dose independently based on these parameters)*

- Type of stimulation (complicated waveforms with drawings):
- Metric to be used (e.g., behavioral, cognitive, EEG, MEP, MRI):
- Product number and model of stimulator used (consider Nr. as encoded in case of multiple stimulators available):
- Stimulation intensity (peak-to-baseline):
- Stimulation duration:

Duration of ramping

Fragmented stimulation (interval duration)

- Type and number of electrodes:
- Electrode positions:
- Electrode polarities in case of tDCS:
- Position of cable fixation at electrode:
- Electrode shape:

target electrode:

return electrode:

- Electrode size:

target electrode:

return electrode:

- Electrode impedance:

target electrode:

return electrode:

- Method of allocation of electrode position (neuronavigation, MEP hot spot, modeling etc.):
- Electrode-skin interface (any skin preparation steps):
- Type of fixation:

saline (molarity?), in case of cream, brand:

Other factors to be considered

- Tasks/status during stimulation (if any): o Not specified or regulated

Specified/regulated: details

- Day time of the experiment (from – to):
- Attention (level of arousal)

1. before stimulation:
2. during stimulation (optimal results expected with relaxation, not during arousal or sleepiness):
3. after stimulation:
4. Number of hours in sleep during the last night:

- Prior motor activity (i.e. cycling before stimulation, if yes, please define the duration):
- Prior rest (sleep) before stimulation:
- Duration of the whole experiment including preparation:
- Number of years in education (of interest in special, e.g. in cognitive studies):
- Additional comments

#### Reading tasks

A behavioural pre-test was administered to 20 typically developing readers (10 children and adolescents: 5 females; M = 12.03 yrs, SD = 1.38 yrs; 10 young adults: 9 females; M = 27.10 yrs, SD = 2.38 yrs). Each participant had to read aloud as rapid and accurate as possible the following reading tasks:

-10 texts of over 400 syllables long (TEXT);

-9 lists of 20 high frequency words (HF - 10 trisyllabic and 10 bisyllabic);

-9 lists of 20 low frequency words (LF - 10 trisyllabic and 10 bisyllabic);

-9 lists of 20 non-words (NW - 10 trisyllabic and 10 bisyllabic) created by rearranging the character string of real word items.

TEXT was written with Time New Roman font, size 13, single-spaced, on a white sheet of A4 paper. HF, LF, NW were arranged in 20-items columns, written with Times New Roman font, size 13, single-spaced, on a white sheet of A4 paper. TEXT derived from an Italian novel (Calvino, 1963). Items in HF list and LF list were matched for Italian written word frequency, number of letters and syllables, bigram frequency (according to CoLFIS, http://www. istc.cnr.it/material/database/colfis) and mean onset reaction time (Barca et al., 2002).

Considering speed, the total time (in terms of seconds) taken to read HF, LF and NW was measured. For TEXT, reading speed was calculated by dividing the total number of syllables spoken by the total time (in terms of seconds) for reading completion and multiplied by 100.

Considering accuracy, for TEXT an error point was assigned in presence of substitution, omission, and/or addition of syllables. A 0.5 error was assigned in case of auto-correction during reading. The percentage of accuracy was considered, calculated by multiplying the ratio of the number of correctly read words to the total number of words presented by 100. For the remaining tasks (HF, LF, NW), an error point was assigned in presence of substitution, omission, and/or addition of syllables, while auto-corrections during reading was not treated as errors. The number of errors was considered.

The accuracy and reading speed of each set of stimuli were compared, and the following equivalent stimuli were selected (see Table S1 for the means (SDs)):

- out of 10 versions, 6 TEXT [Accuracy: F(5, 95) = 0.69, *p* = 0.63, η_p_^2^ = 0.03; Speed: F(5, 95) = 1.80, *p* = 0.12, η_p_^2^ = 0.09];

- out of 9 versions, 7 lists of HF [Accuracy: not possible to perform due to the absence of minimum variance in the data; Speed: F(6, 11) = 1.04, *p* = 0.40, η_p_^2^ = 0.05];

- out of 9 versions, 6 lists of LF [Accuracy: F(5, 95) = 1.15, *p* = 0.34, η_p_^2^ = 0.06; Speed: F(5, 95) = 0.93, *p* = 0.46, η_p_^2^ = 0.05];

- out of 9 versions, 8 lists of NW [Accuracy: F(7, 13) = 0.47, *p* = 0.85, η_p_^2^ = 0.02; Speed: F(7, 13) = 0.77, *p* = 0.61, η_p_^2^ = 0.04].

Table S1. Means (SDs) of reading accuracy and speed for each version of the selected set of stimuli (TEXT, HF, LF, and NW).

| Reading Tasks | | #1 | #2 | #3 | #4 | #5 | #6 | #7 | #8 |
| --- | --- | --- | --- | --- | --- | --- | --- | --- | --- |
| TEXT | Accuracy^a^ | 99.49  (0.54) | 99.39  (0.79) | 99.51  (0.48) | 99.42  (0.49) | 99.33  (0.74) | 99.29  (0.79) | - | - |
|  | Speed^b^ | 5.28  (1.31) | 5.15  (1.10) | 5.38  (1.03) | 5.43  (1.03) | 5.50  (1.01) | 5.41  (1.03) | - | - |
| HF | Accuracy^c^ | 0.05  (0.22) | 00.00  (00.00) | 0.10  (0.45) | 0.20  (0.52) | 0.20  (0.52) | 0.20  (0.41) | 0.05  (0.22) | - |
|  | Speed^d^ | 15.85  (3.12) | 16.10  (3.58) | 15.55  (2.93) | 15.55  (3.44) | 16.40  (4.03) | 16.15  (3.23) | 15.35  (3.15) | - |
| LF | Accuracy^c^ | 0.15  (0.37) | 0.35  (0.67) | 0.20  (0.52) | 0.35  (0.59) | 0.45  (0.60) | 0.45  (0.89) | - | - |
|  | Speed^d^ | 16.80  (3.09) | 17.50  (4.26) | 16.65  (3.41) | 17.25  (3.37) | 17.25  (3.85) | 17.40  (4.36) | - | - |
| NW | Accuracy^c^ | 1.00  (1.21) | 0.85  (1.31) | 1.05  (1.23) | 1.25  (1.58) | 1.20  (1.06) | 1.00  (1.45) | 1.15  (1.53) | 1.05  (1.32) |
|  | Speed^d^ | 29.45  (7.88) | 30.40  (8.62) | 29.90  (7.16) | 31.00  (8.12) | 29.60  (7.98) | 29.25  (7.13) | 29.85  (7.55) | 29.90  (7.82) |
| ^a^ Percentage (%) of accuracy, calculated as accuracy/total number of words x 100; ^b^ Syllables/seconds x 100; ^c^ Number of errors; ^d^ Seconds. HF, High-Frequency words; LF, Low-Frequency words; NW, Non-words. | | | | | | | | |  |

#### Safety and tolerability questionnaire

**Questionnaire of sensations related to transcranial electrical stimulation (TES)**

*(To be filled in by the participants and by the investigator)*

**Investigator:**

**Participant name/code:** ___________________________________**Date:** ____/____/____

**Experiment/Treatment:** ______________________________

**No stimulation experienced before** [ ] **Experienced** [ ]

**# of stimulation sessions before:** ………………….

**Type of electrical stimulation used here** _______________**Intensity** __________mA (if known)

Electrodes dimensions: anode (if known)___*___ cathode (if known) ___*___ (shape_________) other________________

**Participant:**

Did you experience any discomfort during the electrical stimulation? Please indicate the degree of intensity of your discomfort accordingly with the following scale:

- **None**: I did not feel the sensation addressed
- **Mild**: I mildly felt the sensation addressed
- **Moderate**: I felt the sensation addressed
- **Strong**: I felt the sensation addressed to a considerable degree

| *During stimulation session, I felt* | | | | |
| --- | --- | --- | --- | --- |
|  | *None* | *Mild* | *Moderate* | *Strong* |
| Headache | [ ] | [ ] | [ ] | [ ] |
| Neck pain | [ ] | [ ] | [ ] | [ ] |
| Scalp pain | [ ] | [ ] | [ ] | [ ] |
| Tingling | [ ] | [ ] | [ ] | [ ] |
| itching | [ ] | [ ] | [ ] | [ ] |
| Burning sensation | [ ] | [ ] | [ ] | [ ] |
| Skin redness | [ ] | [ ] | [ ] | [ ] |
| Drowsiness | [ ] | [ ] | [ ] | [ ] |
| Concentration difficulties | [ ] | [ ] | [ ] | [ ] |
| Severe mood changes | [ ] | [ ] | [ ] | [ ] |
| Other:_________ | [ ] | [ ] | [ ] | [ ] |

**In case of perceived sensations, when did it begin?**

[	] At the beginning	[	] At approximately in the middle	[	] Towards the end of the stimulation

**Duration (multiple options allowed)**

[	] Only initially	[	] It stopped in the middle of the block	[	] It stopped at the end of the block

**How much did these sensations affect your general state?**

[	] Not at all	[	] Slightly	[	] Considerably	[	] Much	[	] Very much

**Location of sensations:**

[	] Diffuse	[	] Localized	[	] Closed to the electrode, (which one?)____________ [	] Other

If you would like to provide more details, please briefly describe the experimented sensations in relation to the “Other” or “Fatigue” or…. Response:

**To be administered at the end of each stimulation session:**

Do you believe that you received a real or a placebo stimulation?

[	] Real	[	] Placebo	[	] I do not know

**Investigator:**

Please report any adverse event/problem that occurred and rate the event/problem on a scale from 0 to 3 as previously described.

________________________________________________________________________________________________________________________________________________________________________________________________________________________________________________________________________________________________________________________________

Additional comments:

________________________________________________________________________________________________________________________________________________________________________________________________________________________________________________________________________________________________________________________________
